# LENZILUMAB EFFICACY AND SAFETY IN NEWLY HOSPITALIZED COVID-19 SUBJECTS: RESULTS FROM THE LIVE-AIR PHASE 3 RANDOMIZED DOUBLE-BLIND PLACEBO-CONTROLLED TRIAL

**DOI:** 10.1101/2021.05.01.21256470

**Authors:** Zelalem Temesgen, Charles D. Burger, Jason Baker, Christopher Polk, Claudia Libertin, Colleen Kelley, Vincent C. Marconi, Robert Orenstein, Cameron Durrant, Dale Chappell, Omar Ahmed, Gabrielle Chappell, Andrew D. Badley

## Abstract

**BACKGROUND:** Severe COVID-19 pneumonia results from a hyperinflammatory immune response (cytokine storm, CS), characterized by GM-CSF mediated activation and trafficking of myeloid cells, leading to elevation of downstream inflammatory chemokines (MCP-1, IL-8, IP-10), cytokines (IL-6, IL-1), and other markers of systemic inflammation (CRP, D-dimer, ferritin). CS leads to fever, hypotension, coagulopathy, respiratory failure, ARDS, and death. Lenzilumab is a novel Humaneered^®^ anti-human GM-CSF monoclonal antibody that directly binds GM-CSF and prevents signaling through its receptor. The LIVE-AIR Phase 3 randomized, double-blind, placebo-controlled trial investigated the efficacy and safety of lenzilumab to assess the potential for lenzilumab to improve the likelihood of ventilator-free survival (referred to herein as survival without ventilation, SWOV), beyond standard supportive care, in hospitalized subjects with severe COVID-19.

**METHODS:** Subjects with COVID-19 (n=520), ≥18 years, and ≤94% oxygen saturation on room air and/or requiring supplemental oxygen, but not invasive mechanical ventilation, were randomized to receive lenzilumab (600 mg, n=261) or placebo (n=259) via three intravenous infusions administered 8 hours apart. Subjects were followed through Day 28 following treatment.

**RESULTS:** Baseline demographics were comparable between the two treatment groups: male, 64.7%; mean age, 60.5 years; mean BMI, 32.5 kg/m^2^; mean CRP, 98.36 mg/L; CRP was <150 mg/L in 77.9% of subjects. The most common comorbidities were obesity (55.1%), diabetes (53.4%), chronic kidney disease (14.0%), and coronary artery disease (13.6%). Subjects received steroids (93.7%), remdesivir (72.4%), or both (69.1%). Lenzilumab improved the likelihood of SWOV by 54% in the mITT population (HR: 1.54; 95%CI: 1.02-2.31, p=0.041) and by 90% in the ITT population (HR: 1.90; 1.02-3.52, nominal p=0.043) compared to placebo. SWOV also relatively improved by 92% in subjects who received both corticosteroids and remdesivir (1.92; 1.20-3.07, nominal p=0.0067); by 2.96-fold in subjects with CRP<150 mg/L and age <85 years (2.96; 1.63–5.37, nominal p=0.0003); and by 88% in subjects hospitalized ≤2 days prior to randomization (1.88; 1.13-3.12, nominal p=0.015). Survival was improved by 2.17-fold in subjects with CRP<150 mg/L and age <85 years (2.17; 1.04-4.54, nominal p=0.040).

**CONCLUSION:** Lenzilumab significantly improved SWOV in hospitalized, hypoxic subjects with COVID-19 pneumonia over and above treatment with remdesivir and/or corticosteroids. Subjects with CRP<150 mg/L and age <85 years demonstrated an improvement in survival and had the greatest benefit from lenzilumab. NCT04351152

## INTRODUCTION

The clinical manifestations of coronavirus disease 2019 (COVID-19) range from asymptomatic disease to critical illness, acute respiratory disease (ARDS), and death. These pathologies result from the viral-induced hyperinflammatory immune response, or cytokine storm (CS). CS in COVID-19 is similar to that resulting from chimeric antigen receptor T-cell therapy (CAR-T), hemophagocytic lymphohistiocytosis (HLH), macrophage activation syndrome (MAS), and acute graft-versus-host disease (GvHD)^1^; where CS intensity is directly correlated with the severity of tissue injury, neurological manifestations, and adverse clinical outcomes.^2-6^ CS is characterized by granulocyte-macrophage colony-stimulating factor (GM-CSF)-mediated activation and trafficking of myeloid cells^7^, leading to elevations of downstream inflammatory chemokines (MCP-1, IL-8, IP-10), cytokines (IL-6, IL-1), and other markers of systemic inflammation (CRP, D-dimer, ferritin).

In COVID-19, high levels of GM-CSF-secreting T-cells have been associated with disease severity, myeloid cell trafficking to the lungs, and intensive care unit (ICU) admission.^8-10^ Elevation of circulating GM-CSF in subjects with emerging hyperinflammation four days after symptom onset differentiated mild/moderate from severe disease.^10^ Since GM-CSF is produced by activated T-cells in tissue microenvironments and is bound by extracellular matrix and receptors, its detection in the serum indicates significantly elevated tissue levels that orchestrate local and subsequently, systemic hyperinflammatory activity.

CRP levels greater than 100 mg/L upon hospital admission are one of the strongest predictors of critical illness, even more so than age greater than 75 years or other comorbid conditions associated with COVID-19.^11,12^ Subjects with CRP above 150 mg/L are defined as experiencing COVID-19-associated hyperinflammation (COV-HI) and are at risk of escalated respiratory support or death^13^. CRP is a readily available and cost-effective biomarker that can be used to delineate the state of COV-HI, help to identify subjects with emerging CS, and predict the need for early intervention to prevent or reduce progression to IMV and intensive care.^14,15^

Lenzilumab is a novel Humaneered^®^ anti-human GM-CSF monoclonal antibody that directly binds GM-CSF and prevents signaling through its receptor. It has high binding affinity (25 pM) for glycosylated human GM-CSF and a slow off-rate.^16^ Lenzilumab has more than 94% specificity to the human germline to minimize immunogenicity, is highly soluble at a pH of 7.0 to minimize potential for infusion reactions, and in an extensive pre-clinical toxicology program demonstrates a No Observed Adverse Effect Level (NOAEL) of ≥100 mg/kg/week (data on file). No treatment emergent serious adverse events (SAEs) or Suspected Unexpected Serious Adverse Reactions (SUSARSs) have been reported across any of the four completed clinical development studies (data on file). Lenzilumab can be dosed in a regimen designed to achieve therapeutic concentrations to neutralize GM-CSF in both serum and lung tissue. In a matched case-cohort study of high-risk COVID-19 subjects with severe pneumonia; lenzilumab was associated with a significantly shorter time to clinical improvement, defined as at least a 2-point improvement in an 8-point ordinal scale; and an incidence of invasive mechanical ventilation (IMV) or death of 8% compared with 40% (p=0.07) in control patients.^17,18^

LIVE-AIR, a phase 3 randomized, double-blind, placebo-controlled clinical trial was designed to demonstrate that early intervention with lenzilumab, in newly hospitalized COVID-19 subjects who require supplemental oxygen but have not progressed to IMV, improves the likelihood of survival without ventilation (SWOV, sometimes referred to as ventilator-free survival) beyond that provided by supportive care with corticosteroids and remdesivir. Herein are reported the efficacy safety results following 28 days of follow-up.

## METHODS

### Trial Design

The LIVE-AIR trial (NCT04351152) was a phase 3 randomized, double-blind, placebo-controlled study in 520 hospitalized subjects with severe COVID-19 pneumonia across 29 sites in the U.S. and Brazil. The study was conducted in accordance with the Good Clinical Practice guidelines of the International Council for Harmonization E6 and the principles of the Declaration of Helsinki. The protocol was approved by the central/local institutional review board or ethics committee at each site.

All subjects provided written informed consent. Enrolled participants were randomized 1:1 to receive lenzilumab or matched placebo in addition to current standard treatment per institutional guidelines at each site. Subjects were stratified at randomization by age (≤ 65 vs. >65 years) and disease severity (oxygen saturation [SpO_2_]<94% on room air or requiring low-flow supplemental oxygen vs. requirement for high-flow oxygen delivery device or non-invasive positive pressure ventilation [NPPV] or multi-organ dysfunction/failure or shock).

Following screening and baseline measures, participants were administered either lenzilumab or placebo saline beginning at Day 0 within 12 hours from the time of randomization in addition to standard care in accordance with institutional site treatment guidelines. All subjects were monitored at screening, baseline just prior to administration of study drug on Day 0, and at least daily while hospitalized according to the schedule of assessments through Day 28. All primary and key secondary endpoints were assessed by Day 28; however, subjects were followed through Day 60. Adverse events subjects were graded using the National Cancer Institute (NCI) Common Terminology Criteria for Adverse Events (CTCAE) version 5.0, which grades events on a scale from 1 (the mildest intensity) to 5 (fatal). A pharmacokinetic analysis of lenzilumab was also pre-specified and will be reported in a subsequent manuscript.

### Subjects

Eligibility criteria for enrollment included subjects at least 18 years of age who provided informed consent, provided by an authorized proxy, if necessary. SARS-CoV-2 infection was virologically confirmed and pneumonia was diagnosed by chest x-ray or computed tomography. Subjects must have been hospitalized with SpO2 ≤ 94% on room air and/or in need of supplemental oxygen in the form of low-flow oxygen, high-flow oxygen, or NPPV.

Subjects were excluded if they required IMV or extracorporeal membrane oxygenation (ECMO), were pregnant or, in the view of the treating investigator, were not expected to survive the following forty-eight hours from the time of randomization. Subjects with a confirmed diagnosis of bacterial pneumonia or other active/uncontrolled fungal or viral infection other than SARS-CoV-2 were also excluded. Women of childbearing potential were eligible if they had a negative urine or serum pregnancy test at screening/baseline and agreed to adequate contraception following their last dose of study drug.

### Treatments

Lenzilumab or matching placebo (0.9% saline for injection) was administered by intravenous infusion beginning at Day 0 within 12 hours of randomization. Three doses of lenzilumab (600 mg, each) or placebo were administered 8 hours apart via a 1 hour IV infusion, in addition to standard supportive care. Standard pre-infusion medications comprised of acetaminophen 500 to 1000 mg PO or IV and diphenhydramine 12.5 to 25 mg IV, or 25 mg PO or equivalent, and were administered approximately 1-hour prior to infusion of study drug.

Allowed treatments, at baseline and throughout the study, included all current COVID-19 treatments: corticosteroids, convalescent plasma, remdesivir or other anti-virals, and/or hydroxychloroquine with or without azithromycin. Disallowed medications were FDA-approved monoclonal antibodies targeting IL-6 or IL-1, kinase inhibitors, and COVID-19 neutralizing monoclonal antibodies if used within 8 weeks prior to randomization. Sargramostim as well as other investigational therapies to treat COVID-19 were not permitted.

### Outcomes Measures

Assessment of COVID-19 associated pneumonia for development of adult respiratory distress syndrome (ARDS) was conducted during the hospital stay through day 28. This included symptom-directed physical examination, assessment of clinical status using an 8-point ordinal scale adapted (1 = death, 8 = no hospitalization, no limitations) from the NIH-sponsored Adaptive COVID-19 Treatment Trial (ACTT, NCT 04280705)^19^, vital signs, laboratory tests, use of IMV or ECMO, non-invasive ventilation (or high-flow oxygen devices), and low-flow supplemental oxygen, ICU admission, assessment of key cytokines, and adverse events.

The primary efficacy endpoint was a composite endpoint of ventilator-free survival also referred herein as survival without ventilation (SWOV), by Day 28. Failure of the primary endpoint was defined as mortality or the requirement for IMV. SWOV is a robust composite endpoint that is less prone to favor treatments with discordant effects on survival and days free of ventilation^20^ while avoiding the need for sample sizes approaching those of mortality trials to enable timely availability of study results. Key secondary endpoints included the proportion of IMV, ECMO or death, mortality, and Time to Recovery based on the 8 point clinical status scale.

### Sample Size Estimation

The sample size estimate was based on the event rates in similar patient populations from other published studies.^21-23^ The event rate of subjects who required IMV or died by Day 28 in the placebo group was estimated to be approximately 25%, and the event rate in the lenzilumab treatment group was approximated as 15% resulting in a HR of 0.565.^24^ Using a Cox proportional hazard model to test for inequality of the HR, a total of 100 events were calculated as needed to provide 81.47% power to detect a difference using a two-sided alpha of 5.0% at the final analysis and assuming a fixed follow-up of 28 days. Therefore, approximately 516 subjects (258 subjects in each treatment group) were to be enrolled to observe the 100 targeted events.

### Statistical Analysis

The primary endpoint was the difference between lenzilumab treatment and placebo treatment in SWOV through 28 days following treatment in the modified intent to treat population (mITT) who received at least one dose of investigational treatment under the documented supervision of the principal investigator or sub-investigator. This population was defined as the primary analysis and used a Cox proportional hazard model (HR: lenzilumab relative to placebo) accounting for the stratification variables (i.e., age and disease severity) supplemented by a display of K-M curves in each treatment group. The Cox proportional hazard model included the time to first event as the dependent variable (1=IMV use or death, 0=alive with no IMV use); treatment (covariate); and strata (covariates). Where data were non-proportional based on a Chi-squared test proposed by Grambsch and Therneau with a global p-value <0.05, a weighted Cox proportional hazard regression model was used to correct for non-proportionality. For sensitivity and exploratory analyses of the primary endpoint, step-wise addition of all possible two-way interactions between the three covariates was considered. The model with the best fit (lowest AIC value) was selected. Subjects who were alive and did not get placed on IMV were right censored at the date of the last non-missing assessment on or prior to Day 28. The primary analyses were performed in the mITT and ITT populations, respectively, (1) among subjects overall and subjects who (2) had CRP<150 mg/L at screening or if no screening value on day one (baseline) and age <85, (3) received remdesivir and/or any corticosteroid, and (4) randomized ≤2 days from hospitalization and >2 days from hospitalization. The cut-off for CRP was based on the COVID-19-associated hyperinflammation phenotype (COV-HI).^13^ The age cut-off was an original protocol exclusion that was removed in later versions.

For each secondary endpoint, the proportion of subjects that had the event was calculated by treatment group. An odds ratio was calculated for the composite endpoint of the first incident IMV, ECMO or death using logistic regression and including baseline age group and disease category as covariates. Hazard ratios were calculated for each of time to death and time to recovery, separately, as described above. For time to recovery, deaths were censored at Day 28. Subjects who were alive yet did not recover were right censored at the date of the last non-missing assessment of the 8-point clinical status ordinal scale on or prior to Day 28. Last, the proportion of subjects who had CTCAE grade ≥3 treatment-emergent serious adverse events were quantified for each randomization group by system organ class. All data reported herein are reported through Day 28.

Safety was assessed on study drug received, regardless of assignment group. Eight randomized subjects were never treated and were therefore excluded from the safety analysis but were included in the ITT analyses. One subject, randomized to placebo, received lenzilumab in error and was included in the safety analysis of lenzilumab and in the mITT efficacy analysis of placebo.

## RESULTS

### Subjects

Five hundred, twenty-eight subjects were screened, of whom 520 were randomized and included in the ITT population (Figure 1). Forty-one 41 subjects (7.9%) were excluded from the mITT population. Of these, 22 were from two sites in Brazil (12 lenzilumab; 10 placebo) that joined the final stage of the study and experienced documented limitations to access of basic supportive COVID-19 care. These sites experienced an unexpected surge in COVID-19 hospitalizations which affected their ability to provide high-flow oxygen devices, which resulted in a disproportionate increase in the escalation of care from low-flow supplemental oxygen directly to IMV. Subjects from these sites were excluded while the study remained blinded. The mITT population was 236 and 243 subjects in the lenzilumab and placebo groups, respectively: representing 90% and 94% of each randomized population. Despite the challenges, such as those listed above, of conducting a study during a pandemic, the study experienced a loss to follow-up of <2% in each arm and had robust monitoring that yielded 100% source data verification and adherence to good clinical practices.

**Figure 1.**
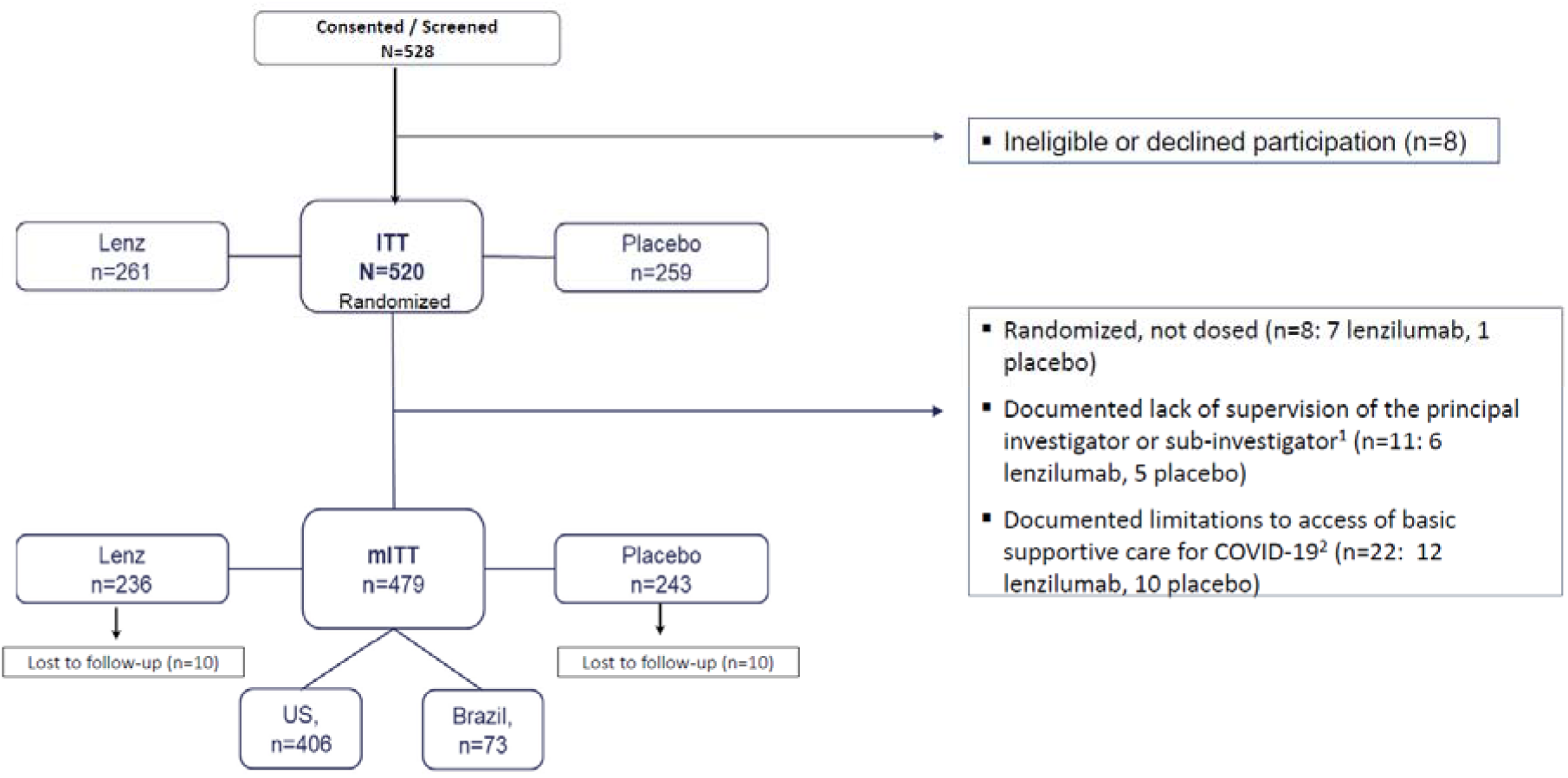
Enrollment and Randomization.

Baseline characteristics were well-balanced between groups (Table 1). Approximately two-thirds of the subjects were male with a mean age of 60.5 years. The subjects were of diverse racial and ethnic backgrounds with 56.8% self-reported as minority or other and 43.2% reporting as white, which is consistent with real-world demographics of hospitalized subjects with COVID-19. At randomization, 40.5% of subjects were ordinal score 3 (high-flow oxygen or NPPV) and 59.5% were ordinal score 4 (low-flow supplemental oxygen) or 5 (SpO2<94% on room air) on the adapted 8-point ordinal scale. Obesity was the most comorbidity followed by diabetes, chronic kidney disease, and coronary artery disease. Subject received corticosteroids (93.7%), remdesivir (72.4%) or both (69.1%) while also receiving study drug. Subjects were hospitalized a median of 2 days prior to randomization. Eighty-five percent of the subjects were enrolled in the US.

**Table 1.** Baseline Characteristics.

|  |  | **Lenzilumab**<br>**(n=236)** | **Placebo**<br>**(n=243)** | **Total**<br>**(n=479)** |
| --- | --- | --- | --- | --- |
| Gender |  |  |  |  |
|  | Male (%) | 64.8 | 64.6 | 64.7 |
| Age |  |  |  |  |
|  | Mean (SD) | 60.5 (13.5) | 60.5 (14.3) | 60.5 (13.9) |
|  | Median (min-max) | 62.0 (28-98) | 62.0 (22-96) | 62.0 (22-98) |
|  | <65 (%) | 60.2 | 58.4 | 59.3 |
|  | ≥ 65 (%) | 39.8 | 41.6 | 40.7 |
|  | >80 (%) | 7.6 | 5.3 | 6.5 |
| BMI |  |  |  |  |
|  | Mean (SD) | 33.1 (8.4) | 31.0 (7.9) | 32.5 (8.2) |
|  | ≥30 Kg/m<sup>2</sup> (%) | 57.6 | 52.7 | 55.1 |
| Race (%) |  |  |  |  |
|  | American Indian | 1.1 | 0.0 | 5.0 |
|  | Asian | 2.7 | 1.2 | 1.9 |
|  | Black | 10.3 | 13.7 | 8.9 |
|  | Hispanic | 35.2 | 42.0 | 38.6 |
|  | White | 45.0 | 41.6 | 43.2 |
|  | Other | 5.2 | 6.2 | 5.8 |
| Region (%) |  |  |  |  |
|  | US | 86.0 | 84.8 | 85.4 |
|  | Brazil | 14.0 | 15.2 | 14.6 |
| Supplemental Oxygen |  |  |  |  |
|  | Room Air or Low-Flow<br>Oxygen | 61.9 | 57.2 | 59.5 |
|  | High Flow Oxygen or NPPV | 38.1 | 42.8 | 40.5 |
| CRP |  |  |  |  |
|  | Mean (SD) | 100.95 (80.14) | 95.88 (71.01) | 98.36 (75.57) |
|  | CRP <150 mg/L (%) | 75.8 | 79.9 | 77.9 |
|  | CRP >150 mg/L (%) | 24.2 | 20.1 | 22.1 |
| Co-Morbidity (%) |  |  |  |  |
|  | *Cardiovascular* |  |  |  |
|  | Congestive Heart Failure | 12.1 | 10.3 | 11.7 |
|  | Coronary Artery Disease | 14.8 | 12.3 | 13.6 |
|  | Diabetes | 50.8 | 56.0 | 53.4 |
|  | Chronic Liver Disease | 4.2 | 5.8 | 5.0 |
|  | Chronic Kidney Disease | 14.4 | 13.6 | 14.0 |
|  | *Respiratory* |  |  |  |
|  | Asthma | 13.6 | 7.8 | 10.6 |
|  | Interstitial Pulmonary Fibrosis | 1.3 | 0.4 | 0.8 |
|  | COPD | 7.6 | 7.0 | 7.3 |
| Treatments (%) |  |  |  |  |
|  | Remdesivir | 72.1 | 72.8 | 72.4 |
|  | Corticosteroids | 93.7 | 93.8 | 93.7 |
|  | Remdesivir & Corticosteroids | 69.1 | 69.1 | 69.1 |

### Primary Outcome

The study achieved its pre-specified primary endpoint. In the mITT population, the Cox proportional hazard model estimated a 54% relative improvement in the likelihood of subjects treated with lenzilumab achieving SWOV compared to the placebo group (HR: 1.54; 95%CI: 1.02-2.31, p=0.041; Table 2, Figure 2a). Failure to achieve SWOV occurred in 15.6% and 22.1% of subjects treated with lenzilumab or placebo, respectively (Table 2). Separation of the survival curves occurred as early as 3 days following treatment (Figure 2a), continued to increase through approximately Day 10, and then was maintained for the duration of the 28-day observation period. In the ITT population, lenzilumab improved the likelihood of SWOV by 1.90-fold (nominal p=0.043; Table 2).

**Table 2.**
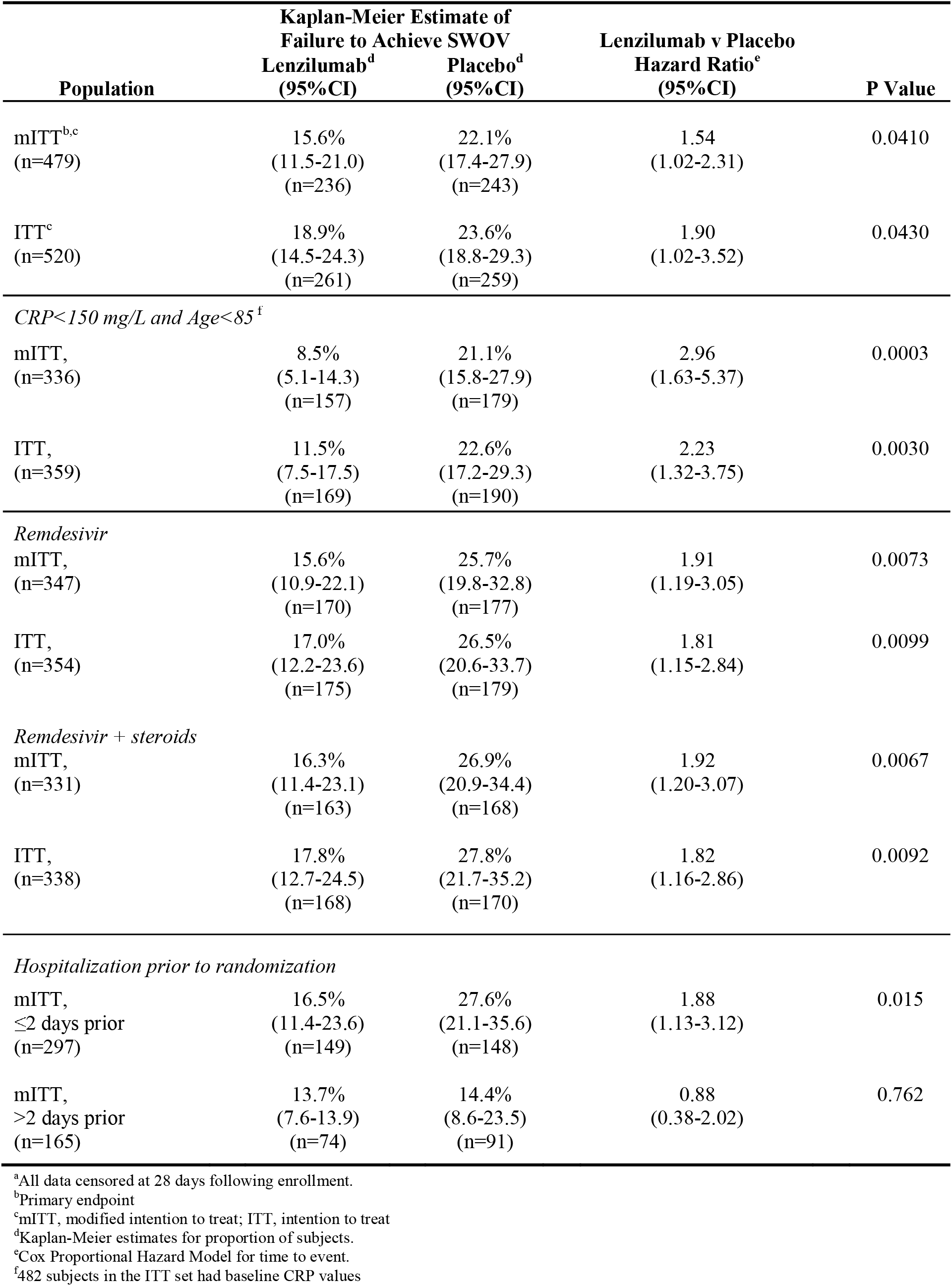
Survival Without Ventilation.

| Population | Kaplan-Meier Estimate of<br>Failure to Achieve SWOV |  | Lenzilumab v Placebo<br>Hazard Ratio<sup>e</sup><br>(95% CI) | P Value |
| --- | --- | --- | --- | --- |
|  | Lenzilumab<sup>d</sup><br>(95% CI) | Placebo<sup>d</sup><br>(95% CI) |  |  |
| mITT<sup>b,c</sup><br>(n=479) | 15.6%<br>(11.5-21.0)<br>(n=236) | 22.1%<br>(17.4-27.9)<br>(n=243) | 1.54<br>(1.02-2.31) | 0.0410 |
| ITT<sup>c</sup><br>(n=520) | 18.9%<br>(14.5-24.3)<br>(n=261) | 23.6%<br>(18.8-29.3)<br>(n=259) | 1.90<br>(1.02-3.52) | 0.0430 |
| *CRP < 150 mg/L and Age < 85<sup>f</sup>* |  |  |  |  |
| mITT,<br>(n=336) | 8.5%<br>(5.1-14.3)<br>(n=157) | 21.1%<br>(15.8-27.9)<br>(n=179) | 2.96<br>(1.63-5.37) | 0.0003 |
| ITT,<br>(n=359) | 11.5%<br>(7.5-17.5)<br>(n=169) | 22.6%<br>(17.2-29.3)<br>(n=190) | 2.23<br>(1.32-3.75) | 0.0030 |
| *Remdesivir* |  |  |  |  |
| mITT,<br>(n=347) | 15.6%<br>(10.9-22.1)<br>(n=170) | 25.7%<br>(19.8-32.8)<br>(n=177) | 1.91<br>(1.19-3.05) | 0.0073 |
| ITT,<br>(n=354) | 17.0%<br>(12.2-23.6)<br>(n=175) | 26.5%<br>(20.6-33.7)<br>(n=179) | 1.81<br>(1.15-2.84) | 0.0099 |
| *Remdesivir + steroids* |  |  |  |  |
| mITT,<br>(n=331) | 16.3%<br>(11.4-23.1)<br>(n=163) | 26.9%<br>(20.9-34.4)<br>(n=168) | 1.92<br>(1.20-3.07) | 0.0067 |
| ITT,<br>(n=338) | 17.8%<br>(12.7-24.5)<br>(n=168) | 27.8%<br>(21.7-35.2)<br>(n=170) | 1.82<br>(1.16-2.86) | 0.0092 |
| *Hospitalization prior to randomization* |  |  |  |  |
| mITT,<br>≤ 2 days prior<br>(n=297) | 16.5%<br>(11.4-23.6)<br>(n=149) | 27.6%<br>(21.1-35.6)<br>(n=148) | 1.88<br>(1.13-3.12) | 0.015 |
| mITT,<br>> 2 days prior<br>(n=165) | 13.7%<br>(7.6-13.9)<br>(n=74) | 14.4%<br>(8.6-23.5)<br>(n=91) | 0.88<br>(0.38-2.02) | 0.762 |

<sup>a</sup>All data censored at 28 days following enrollment.<sup>b</sup>Primary endpoint<sup>c</sup>mITT, modified intention to treat; ITT, intention to treat<sup>d</sup>Kaplan-Meier estimates for proportion of subjects.<sup>e</sup>Cox Proportional Hazard Model for time to event.<sup>f</sup>482 subjects in the ITT set had baseline CRP values

**Table 3.** Secondary Efficacy Outcomes, mITT^a^.

| **Outcome** | **Lenzilumab** | **Placebo** | **Hazard Ratio**<br>**or Odds Ratio**<br>**(95% CI)** | **P Value** |
| --- | --- | --- | --- | --- |
| *Incidence IMV,*<br>*ECMO or death (%)* | 15.4<br>(11.1-21.0)<br>(n=236) | 21.4<br>(16.3-27.4)<br>(n=243) | 0.67<br>(0.41-1.10) | 0.111 |
| CRP<150 mg/L<br>Age<85<br>(n=336) | 7.2<br>(4.0-12.7)<br>(n=157) | 19.5<br>(13.8-26.9)<br>(n=179) | 0.32<br>(0.15-0.65) | 0.002 |
| *Mortality (%)* | 9.6<br>(6.4-14.2)<br>(n=236) | 13.9<br>(10.1-19.0)<br>(n=243) | 1.38<br>(0.81-2.37) | 0.239 |
| CRP<150 mg/L,<br>Age<85<br>(n=336) | 6.6<br>(3.6-11.9)<br>(n=157) | 13.8<br>(9.5-19.9)<br>(n=179) | 2.17<br>(1.04-4.54) | 0.040 |
| *Time to Recovery*<br>*(median days)* | 8<br>(7-9)<br>(n=236) | 8<br>(7-9)<br>(n=243) | 1.09<br>(0.88-1.35) | 0.43 |
| CRP<150 mg/L,<br>Age<85<br>(n=336) | 7<br>(6-7)<br>(n=157) | 8<br>(7-8)<br>(n=179) | 1.36<br>(1.07-1.73) | 0.012 |

<sup>a</sup>All data censored at 28 days following enrollment.<sup>b</sup>Kaplan-Meier estimates for proportion of subjects. Value represent estimate (95% CI)<sup>c</sup>Cox Proportional Hazard Model for time to event.

**Figure 2.**
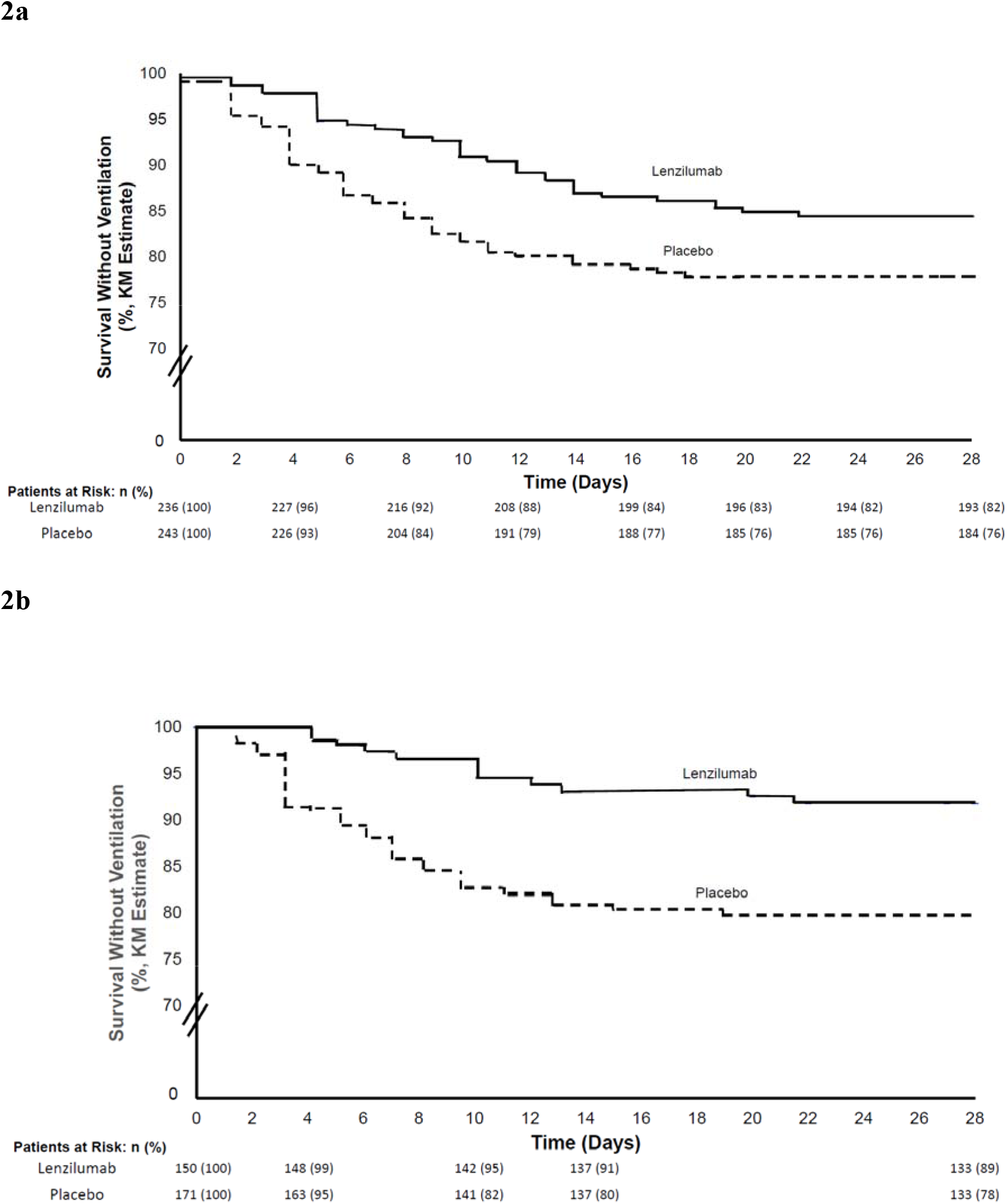
Kaplan Meier plot of Survival Without Ventilation. a. Plot for mITT population. b. Plot for mITT population with baseline CRP<150 mg/L

The primary outcome was substantiated by sensitivity analysis Baseline CRP was obtained in 482 subjects, of which 77.9% had a CRP level of <150 mg/L. In these subjects who were also < 85 years of age, lenzilumab improved the likelihood of SWOV by 2.96-fold (nominal p=0.0003) in the mITT analysis (Table 2, Figure 2b). A second sensitivity analysis on the ITT population with CRP<150 mg/L and <85 years old, lenzilumab improved SWOV by 2.23-fold (nominal p=0.003; Table 2). Additional sensitivity analyses demonstrated the significant improvement of SWOV with lenzilumab treatment over and above any improvement afforded by the combined use of remdesivir plus steroids (n=331) by 1.92-fold (nominal p=0.0067; Table 2). SWOV in subjects treated with lenzilumab was significantly improved by 1.88-fold (nominal p=0.015) when randomization day was no more than the median of two days after hospitalization (Table 2).

Further analysis of CRP<150 mg/L demonstrates a sigmoidal relationship where subjects with these characteristics receive the greatest likelihood of achieving SWOV (Figure3). Lenzilumab decreased CRP levels (Figure 4a), beginning two days after administration and continuing for 28 days, but only in subjects with a baseline CRP<150 mg/L (Figure 4b).

**Figure 3.**
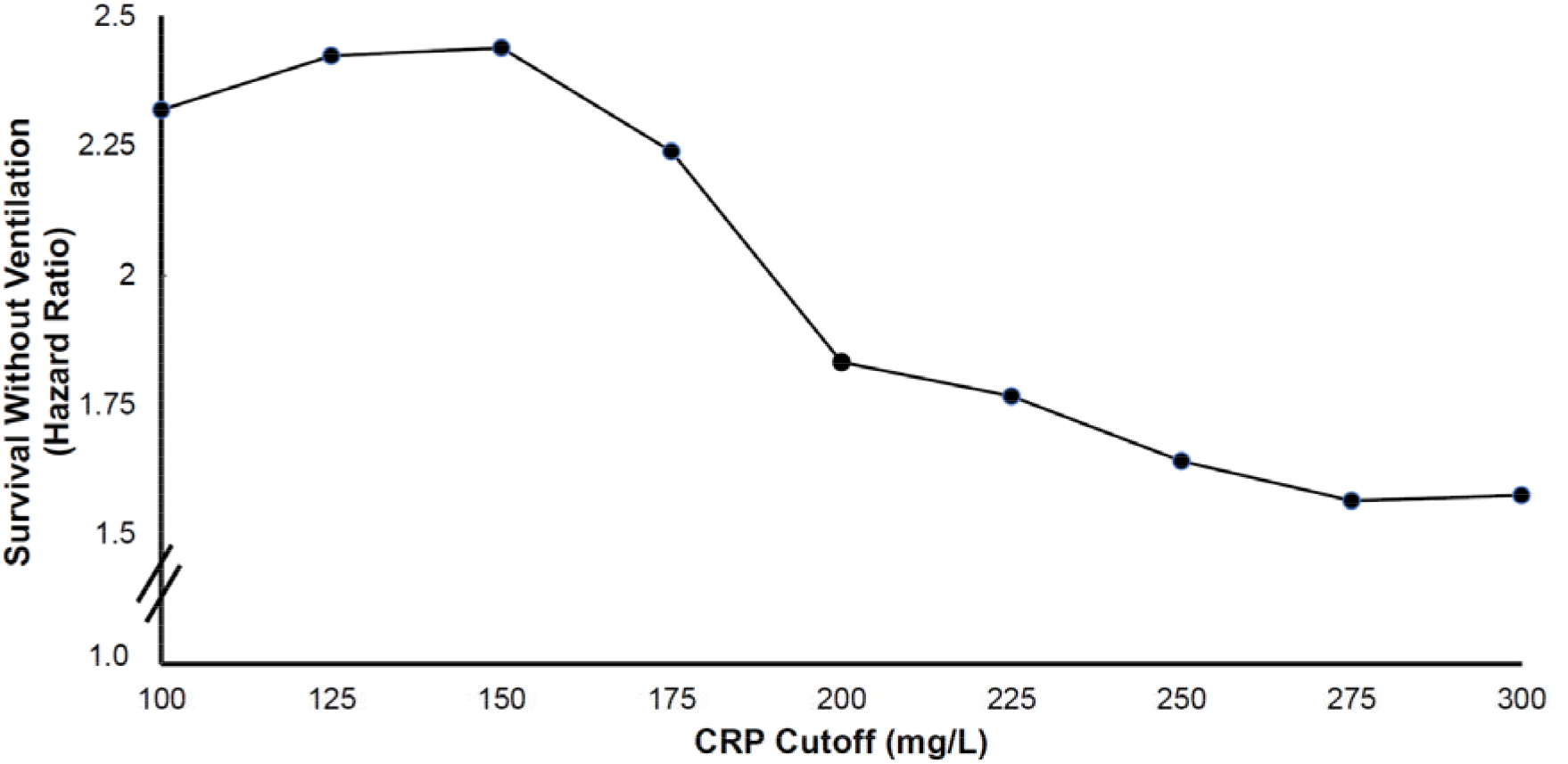
Prediction of Survival Without Ventilation by Level of CRP Cutoff. The hazard ratio for survival without ventilation was calculated for all patients regardless of age with CRP level below the cutoff value.

**Figure 4.**
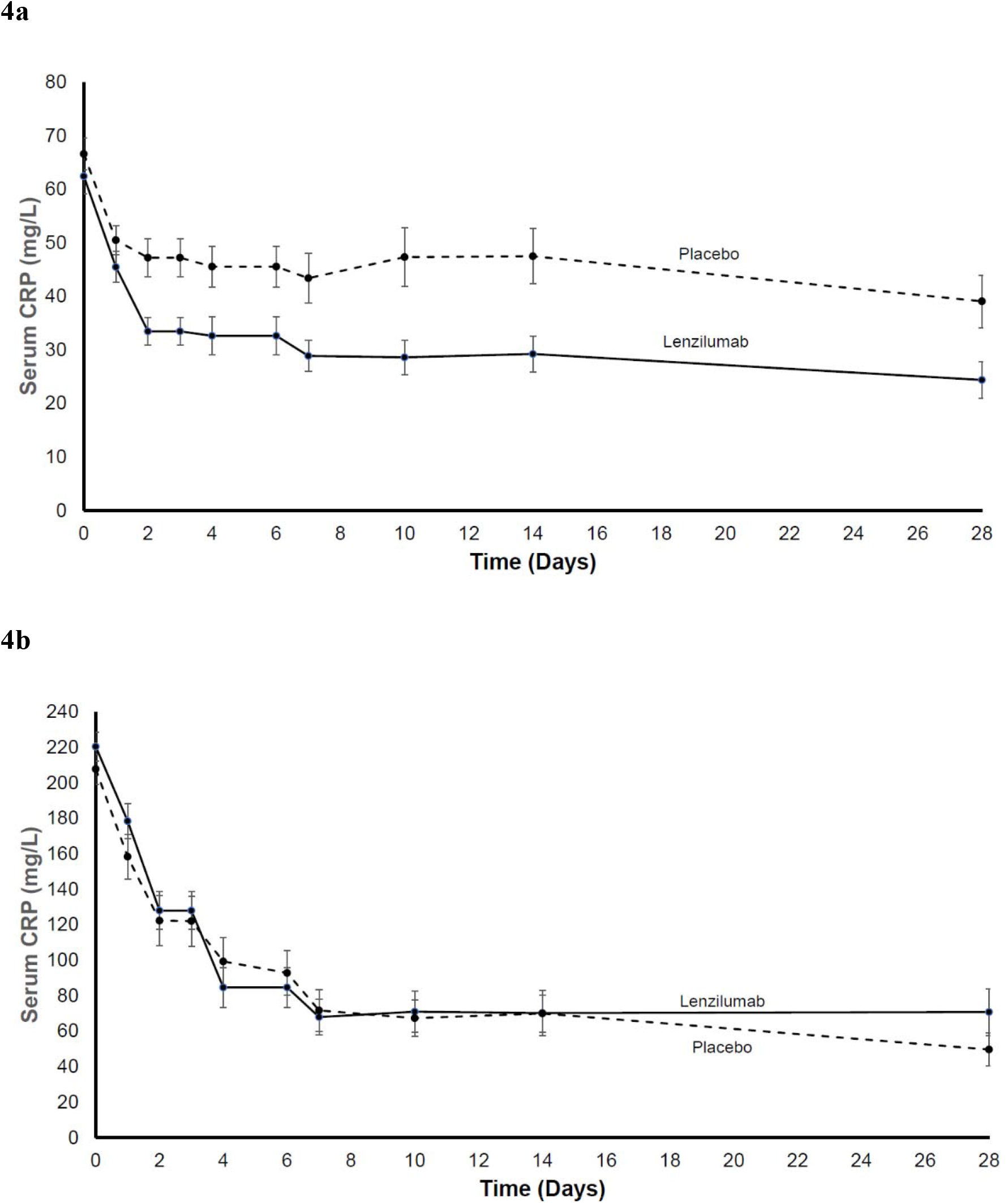
Change in Serum CRP Following Treatment. a. CRP in mITT population with baseline with CRP<150 mg/L. b. CRP in mITT population with baseline CRP≥150 mg/L (with standard error).

### Secondary Outcomes

Incidence of IMV, ECMO or death was 15.4% in the lenzilumab group and 21.4% in placebo (OR 0.67; 95%CI 0.41-1.10), p=ns) but was significantly improved in subjects with CRP<150 mg/L and age<85 (OR 0.32; 95%CI, 0.15-0.65, nominal p=0.002). Mortality was significantly improved by 2.17-fold in subjects with CRP<150 mg/L and age<85 (nominal p=0.040). Similarly, Time to Recovery was not different between groups in the mITT population but was significantly improved by 36% (nominal p=0.012) in the mITT population with CRP<150 mg/L and age<85.

### Safety

In the safety population, adverse events ≥ Grade 3 were reported in 26.7% and 32.7% of the lenzilumab and placebo subjects, respectively (Table 4). Serious adverse events were also similar and reported in 24.7% and 29.6% of the subjects in the lenzilumab and placebo groups, respectively. Lenzilumab, compared with placebo, produced no infusion-related reactions, and no attributable serious adverse events; including, hematologic laboratory abnormalities, liver enzyme abnormalities, pulmonary alveolar proteinosis, or increased incidence of infection.

**Table 4.** Most Common Grade ≥3 Adverse Events (Overall Incidence ≥ 1.0%)

| **System Organ Class**<br>**Preferred Term n (%)** | **Lenzilumab**<br>**N=255** | **Placebo**<br>**N=257** | **Total**<br>**N=512** |
| --- | --- | --- | --- |
| Any AE ≥ Grade 3 | 68 (26.7) | 84 (32.7) | 152 (29.7) |
| Respiratory, thoracic and mediastinal disorders | 64 (25.1) | 71 (27.6) | 135 (26.4) |
| Respiratory failure | 24 (9.4) | 31 (12.1) | 55 (10.7) |
| Acute respiratory failure | 18 (7.1) | 22 (8.6) | 40 (7.8) |
| Hypoxia | 15 (5.9) | 15 (5.8) | 30 (5.9) |
| Pulmonary embolism | 5 (2.0) | 3 (1.2) | 8 (1.6) |
| Acute respiratory distress syndrome | 4 (1.6) | 3 (1.2) | 7 (1.4) |
| Cardiac disorders | 15 (5.9) | 14 (5.4) | 29 (5.7) |
| Cardiac arrest | 8 (3.1) | 4 (1.6) | 12 (2.3) |
| Cardio-respiratory arrest | 3 (1.2) | 4 (1.6) | 7 (1.4) |
| Infections and infestations | 10 (3.9) | 16 (6.2) | 26 (5.1) |
| Septic shock | 5 (2.0) | 9 (3.5) | 14 (2.7) |
| Sepsis | 2 (0.8) | 5 (1.9) | 7 (1.4) |
| Pneumonia bacterial | 0 (0.0) | 6 (2.3) | 6 (1.2) |
| Vascular disorders | 10 (3.9) | 15 (5.8) | 25 (4.9) |
| Shock | 3 (1.2) | 6 (2.3) | 9 (1.8) |
| Hypotension | 2 (0.8) | 5 (1.9) | 7 (1.4) |
| Renal and urinary disorders | 5 (2.0) | 11 (4.3) | 16 (3.1) |
| Acute kidney injury | 5 (2.0) | 8 (3.1) | 13 (2.5) |
| General disorders and administration site conditions | 4 (1.6) | 11 (4.3) | 15 (2.9) |
| Multiple organ dysfunction syndrome | 3 (1.2) | 6 (2.3) | 9 (1.8) |

## DISCUSSION

The LIVE-AIR trial in newly hospitalized COVID-19 patients, who were hypoxic but had not progressed to IMV showed that early lenzilumab treatment improved the relative likelihood of SWOV through Day 28 by 54%. This finding was supported by sensitivity analyses in the ITT population. Further sensitivity analyses showed that the improvement in likelihood of SWOV was over and above treatment with remdesivir and corticosteroids. Additionally, improvement in SWOV was observed in subjects hospitalized for less than 2 days. Finally, improvement in SWOV was most evident in patients with CRP<150 mg/L who are less than 85 years of age. Lenzilumab was associated with similar improvements on key secondary endpoints including; incidence of IMV, ECMO or death, mortality, and time to recovery in those subjects with CRP<150 mg/L and < 85 years. Treatment with lenzilumab was safe and well tolerated with a serious adverse event profile no different from placebo.

Safe and effective therapeutics to treat hospitalized subjects with COVID-19 remains a significant unmet clinical need. The NIH currently recommends against the use of dexamethasone in hospitalized, hypoxic subjects who do not require supplemental oxygen and data regarding the benefit of remdesivir in this population are conflicting and insufficient to recommend for use.^25^ Recommendations supporting the combination of dexamethasone and remdesivir in hospitalized subjects on low-flow supplemental oxygen are based primarily on expert opinion according to the NIH as no randomized controlled studies have evaluated these agents in combination.^25^ The primary population where the use of dexamethasone has proven to significantly improve mortality is in subjects on IMV who are <70 years of age. Data reported on the use of tocilizumab is conflicting; and baricitinib is reserved for those with contraindications to corticosteroid. In LIVE-AIR, early intervention with lenzilumab improved the relative likelihood of SWOV through Day 28 by 54% in hospitalized, racially and ethnically diverse subjects with COVID-19, who had several comorbid conditions and were hypoxic but did not require mechanical ventilation at baseline.

CRP was analyzed as it is a marker of systemic inflammation, routinely used clinically and known to be proportionally elevated with the severity of COVID-19 disease.^11-13^ In this trial, 77% of subjects with an evaluable CRP, had a baseline value <150 mg/L and 74% had a baseline CRP value <150 mg/L and age<85. The CRP analysis clearly demonstrated that CRP less than 150 mg/L was associated with best likelihood of achieving SWOV and clearly differentiated emerging hyperinflammation from full-blown CS. In subjects with CRP<150 mg/L and < 85 years old, lenzilumab improved SWOV by nearly 3-fold and was accompanied by a 2.17-fold improvement in mortality as well as a 36% improvement in time to recovery. Given that CRP is a readily available, routinely measured biomarker of systemic inflammation, with prognostic value affirmed in this and other trials^11,12^, it can clearly be used to clinically identify a phenotype of COVID-19 disease that best responds to lenzilumab treatment.

LIVE-AIR used a composite primary endpoint of SWOV which includes time to death or time to invasive mechanical ventilation. This endpoint has been called “ventilator-free survival” and “alive and ventilator-free”. The nomenclature selected in LIVE-AIR more accurately captures the meaning of the endpoint without confusion by alternate interpretations. Regardless of nomenclature, the composite endpoint is appropriate for an event-based clinical trial of this size to predict a clinically meaningful outcome that equates ventilation with mortality.^20^ It is not confounded by mechanisms that may preferentially affect either ventilation or death.^20^ Lenzilumab’s ability to improve SWOV was more pronounced in combination with current treatments including remdesivir which has demonstrated a 5 day improvement in time to recovery^23^ and dexamethasone which improved survival in subjects needing respiratory support^21,26^ which may indicate a synergistic mechanism of action with the triple combination. This robust improvement in the composite endpoint demonstrates the potential for lenzilumab treatment to result in a true improvement in survival since nearly all subjects that die from COVID-19 have received IMV during their hospital stay. Confirmation of the lenzilumab improvement of survival, and the role of SWOV as a surrogate endpoint, could be confirmed in a large adequately powered mortality trial.

The results with lenzilumab on SWOV demonstrate that impeding GM-CSF signaling early in CS is required to prevent COVID-19 disease progression to IMV or death. This is further supported by an 88% improvement in SWOV in subjects hospitalized < 2 days. The results herein with lenzilumab confirm results from prior clinical trials targeting GM-CSF but are differentiated by patient selection, timing of intervention, and dosing strategy. Mavrilimumab, a monoclonal antibody that acts as a GM-CSF alpha-receptor antagonist demonstrated non-significant trends of 48% improvement in survival without requirement for supplemental oxygen.^27^ Otilimab, a monoclonal antibody targeting circulating GM-CSF in ventilated subjects, was reported in a non-peer-reviewed disclosure of results from its phase 2b COVID-19 study (NCT04376684) to not meet its primary endpoint of improvement in the proportion of COVID-19 subjects who were alive and free of respiratory failure 28 days; however, in pre-planned analysis of subjects aged at least 70 years, which comprised 22% of the entire patient population, 65.1% were alive and free of respiratory failure, compared to 45.9% of subjects who received the standard of care.^28^ The results of all three trials of GM-CSF antagonism suggest the central role of GM-CSF in CS of COVID-19 that has been recently demonstrated through network analysis of cytokine profiles from COVID-19 subjects.^10^ This analysis associated IL-6, CXCL10, but in particular GM-CSF, to markers of endothelial injury and thrombosis early in COVID-19, in severe cases, in hospitalized subjects and those with risk factors for severe disease, and uniquely elevated in fatal COVID-19. The results herein with lenzilumab more completely define the central role of GM-CSF in COVID-19, identified subjects more likely to receive benefit from lenzilumab and underscores the necessity to target GM-CSF early during the emerging hyperinflammatory stage in order to ameliorate CS induced by COVID-19.

The findings of the network analysis and the anti-GM-CSF approaches listed above, including the robust response to lenzilumab particularly in subjects with CRP levels <150 mg/L and age<85, helps explain the limited efficacy of other anti-cytokine approaches that target cytokines downstream of myeloid cell activation in the CS cascade (i.e., IL-6, TNF?, IL-10). Results from clinical trials with anti-IL-6 agents are mixed and mostly negative.^22,29-33^. Targeting single downstream mediators of CS, such as IL-6, may render therapy clinically ineffective as other key, redundant cytokines continue to promote tissue damage or are even compensatory upregulated in the absence of IL-6 signaling. Alternatively, it is possible that targeting IL-6 is more effective later in the disease course as evidenced by tocilizumab wherein benefit appears to be derived in more critically ill subjects recently admitted to the ICU.^22^

In the LIVE-AIR study, lenzilumab was studied as a real-world early intervention in hospitalized, hypoxic subjects not requiring invasive mechanical ventilation; hypothesizing that neutralizing GM-CSF during emerging hyperinflammation, could prevent the consequences of full-blown CS, in subjects at risk but already treated with standard of care. Lenzilumab improved the robust endpoint of SWOV, the effect of which was more pronounced in emerging hyperinflammation than in subjects who were further evolved in their disease course. LIVE-AIR has two important limitations. First, while it demonstrated an improvement in survival in subjects with CRP<150 mg/L and aged <85, the study was not powered to observe a survival benefit, and indeed did not achieve statistical significance on this key secondary endpoint in the mITT population. Second, the observations pertaining to benefits in the population that had CRP<150 mg/L and age <85 were exploratory.

Despite high vaccine efficacy and uptake in some countries, hospitalizations for COVID-19 will continue to occur given the propensity for mutation and escape from protection that occurs through natural immunological selection.^34^ A proportion of those subjects will unfortunately progress to hypoxia and hospitalization. For them, dexamethasone is currently the only treatment approach shown to provide a survival benefit. This study demonstrates that early neutralization of GM-CSF with lenzilumab, a key initiator and orchestrator of CS, in newly hospitalized hypoxic subjects can improve their likelihood of survival without the need for mechanical ventilation and defines a targeted patient population most likely to derive the greatest benefit over and above that of steroids and/or remdesivir.

## Data Availability

The authors confirm that the data supporting the findings of this study are available within the article

## ACKNOWLEDGEMNTS

The LIVE-AIR study team thanks and acknowledges all subjects for their participation in this important research; CTI Clinical Trial & Consulting Services for their support as the clinical research organization managing the operations of the study; RXMD as well as all HGEN003-06 study investigators and their research teams, including but not limited to Drs. Meghan Lewis, Linda Sher, Michael Bowdish, Noah Wald-Dickler, Subarna Biswas, Lydia Lam, Khang Vo, Roy Poblete, May M. Lee, Douglass Hutcheon from the University of Southern California (USC) Keck and LAC Medical Centers; Drs. John Gharbin, Caitlin Moran, Sheetal Kandiah, Valeria Cantos, Paulina Rebolledo, Carlos del Rio, Jeffrey Lennox, Carmen Polito, Paulina Rebolledo, Anandi Sheth, Michael Woodworth from Emory University Medical Center and Grady Memorial Hospital; Dr. Anup Patel from St. Barnabas Medical Center; Dr. Seife Yohannes from MedStar Washington Hospital Center; Drs. Alpesh Amin, Richard Lee, Miki Watanabe, Lanny Hsieh from the University of California-Irvine Medical Center; Drs. Martin Cearras, Amay Parikh, Jason Sniffen, Wilfred Onyia from AdventHealth Orlando; Drs. Michael Boger, Lisa Davidson, Kiran Gajurel, Michael Leonard, Lewis McCurdy, Nestor Quezada, Mindy Sampson, Zainab Shahid, Stephanie Strollo, David Weinrib, Sara Zulfigar from Atrium Health; Drs. Cheryl McDonald, John Hollingsworth, John Burk, Joshua Berg, Daniel Barbaro, Lakshmi Sambathkumar, Stuart McDonald, Obinna Okoye from Texas Health Harris Methodist Hospital Fort Worth; Drs. Juan Pulido, J. Fulton, W. Gill from Baptist Health Research Institute Jacksonville; Drs. Richard Zuckerman, Lionel Lewis from Dartmouth-Hitchcock Medical Center; Dr. Chaitanya Mandapakala from St. Elizabeth Medical Center; Drs. Matthew Robinson, Brian Metzger from St. David’s Medical Center; Drs. Maqsood Alam, Chrisoula Politis from Mercy Medical; Drs. Anne Frosch, Linh Ngo from Hennepin Healthcare; Dr. Fernando Neunschwander from Hospital Vera Cruz in Brazil; Dr. Ricardo Diaz from UNIFESP in Brazil; Dr. Marina Lima from Hospital Dia do Pulmaeo in Brazil; Dr. Daniel Santos from IDOR in Brazil; Dr. Juvencio Jose Furtado from Hospital Heliopolis in Brazil; Fabiano Ramos from Hospital Sao Lucas – PUCRS; Kleber Luz from CPCLIN in Brazil; Felipe Dal Pizzol from SLCSA in Brazil; Adilson Cavalcnate from CEMEC in Brazil; Martti Antilain from CMPC in Brazil;

## REFERENCES

1. Fajgenbaum DC, June CH. Cytokine Storm. N Engl J Med 2020;383:2255–73.

2. Arnaldez FI, O’Day SJ, Drake CG, et al. The Society for Immunotherapy of Cancer perspective on regulation of interleukin-6 signaling in COVID-19-related systemic inflammatory response. J Immunother Cancer 2020;8.

3. Ascierto PA, Fox BA, Urba WJ, et al. Insights from immuno-oncology: the Society for Immunotherapy of Cancer Statement on access to IL-6-targeting therapies for COVID-19. J Immunother Cancer 2020;8.

4. Henry BM, de Oliveira MHS, Benoit S, Plebani M, Lippi G. Hematologic, biochemical and immune biomarker abnormalities associated with severe illness and mortality in coronavirus disease 2019 (COVID-19): a meta-analysis. Clin Chem Lab Med 2020;58:1021–8.

5. Wang W, Liu X, Wu S, et al. Definition and Risks of Cytokine Release Syndrome in 11 Critically Ill COVID-19 Patients With Pneumonia: Analysis of Disease Characteristics. J Infect Dis 2020;222:1444–51.

6. Pensato U, Muccioli L, Cani I, et al. Brain dysfunction in COVID-19 and CAR-T therapy: cytokine storm-associated encephalopathy. Ann Clin Transl Neurol 2021;8:968–79.

7. Xiong Y, Liu Y, Cao L, et al. Transcriptomic characteristics of bronchoalveolar lavage fluid and peripheral blood mononuclear cells in COVID-19 patients. Emerg Microbes Infect 2020;9:761–70.

8. Zhou Y, Fu B, Zheng X, et al. Pathogenic T cells and inflammatory monocytes incite inflammatory storm in severe COVID-19 patients. Natl Sci Rev 2020:nwaa041.

9. Kox M, Waalders NJB, Kooistra EJ, Gerretsen J, Pickkers P. Cytokine Levels in Critically Ill Patients With COVID-19 and Other Conditions. Jama 2020.

10. Thwaites RS, Sanchez Sevilla Uruchurtu A, Siggins MK, et al. Inflammatory profiles across the spectrum of disease reveal a distinct role for GM-CSF in severe COVID-19. Sci Immunol 2021;6.

11. Lavillegrand JR, Garnier M, Spaeth A, et al. Elevated plasma IL-6 and CRP levels are associated with adverse clinical outcomes and death in critically ill SARS-CoV-2 patients: inflammatory response of SARS-CoV-2 patients. Ann Intensive Care 2021;11:9.

12. Petrilli CM, Jones SA, Yang J, et al. Factors associated with hospital admission and critical illness among 5279 people with coronavirus disease 2019 in New York City: prospective cohort study. Bmj 2020;369:m1966.

13. Manson JJ, Crooks C, Naja M, et al. COVID-19-associated hyperinflammation and escalation of patient care: a retrospective longitudinal cohort study. Lancet Rheumatol 2020;2:e594–e602.

14. Hodges G, Pallisgaard J, Schjerning Olsen AM, et al. Association between biomarkers and COVID-19 severity and mortality: a nationwide Danish cohort study. BMJ open 2020;10:e041295.

15. Keddie S, Ziff O, Chou MKL, et al. Laboratory biomarkers associated with COVID-19 severity and management. Clin Immunol 2020;221:108614.

16. Padron E, Painter JS, Kunigal S, et al. GM-CSF-dependent pSTAT5 sensitivity is a feature with therapeutic potential in chronic myelomonocytic leukemia. Blood 2013;121:5068–77.

17. Temesgen Z, Assi M, Shweta FNU, et al. GM-CSF Neutralization With Lenzilumab in Severe COVID-19 Pneumonia: A Case-Cohort Study. Mayo Clin Proc 2020;95:2382–94.

18. Temesgen Z, Assi M, Vergidis P, et al. First Clinical Use of Lenzilumab to Neutralize GM-CSF in Patients with Severe COVID-19 Pneumonia. medRxiv 2020.

19. Natiotnal Institute of Allergy and Infectious Diseases. Adaptive COVID-19 Treatment Trial (ACTT): NCT04280705. 2020. Accessed April 24, 2021, at https://clinicaltrials.gov/ct2/show/record/NCT04280705.)

20. Novack V, Beitler JR, Yitshak-Sade M, et al. Alive and Ventilator Free: A Hierarchical, Composite Outcome for Clinical Trials in the Acute Respiratory Distress Syndrome. Critical care medicine 2020;48:158–66.

21. RECOVERY Collaborative Group, Horby P, Lim WS, et al. Dexamethasone in Hospitalized Patients with Covid-19. N Engl J Med 2021;384:693–704.

22. Horby PW, Pessoa-Amorim G, Peto L, et al. Tocilizumab in patients admitted to hospital with COVID-19 (RECOVERY): preliminary results of a randomised, controlled, open-label, platform trial. medRxiv 2021:2021.02.11.21249258.

23. Beigel JH, Tomashek KM, Dodd LE, et al. Remdesivir for the Treatment of Covid-19 - Final Report. N Engl J Med 2020;383:1813–26.

24. Mehta CR, Pocock SJ. Adaptive increase in sample size when interim results are promising: a practical guide with examples. Stat Med 2011;30:3267–84.

25. National Institutes of Health. Therapeutic Management of Adults With COVID-19. Updated April 21, 2021, Accessed April 27, 2021, at https://www.covid19treatmentguidelines.nih.gov/therapeutic-management/.)

26. Tomazini BM, Maia IS, Cavalcanti AB, et al. Effect of Dexamethasone on Days Alive and Ventilator-Free in Patients With Moderate or Severe Acute Respiratory Distress Syndrome and COVID-19: The CoDEX Randomized Clinical Trial. Jama 2020;324:1307–16.

27. Cremer PC, Abbate A, Hudock K, et al. Mavrilimumab in patients with severe COVID-19 pneumonia and systemic hyperinflammation (MASH-COVID): an investigator initiated, multicentre, double-blind, randomised, placebo-controlled trial. Lancet Rheumatol 2021.

28. Glaxo Smith Kline. GSK announces results evaluating its investigational monoclonal antibody, otilimab, for the treatment of hospitalised adult patients with COVID-19. 2021. Accessed April 21, 2021 at https://www.gsk.com/en-gb/media/press-releases/gsk-announces-otilimab-data-for-treatment-of-covid-19/.)

29. Salvarani C, Dolci G, Massari M, et al. Effect of Tocilizumab vs Standard Care on Clinical Worsening in Patients Hospitalized With COVID-19 Pneumonia: A Randomized Clinical Trial. JAMA Intern Med 2021;181:24–31.

30. Rosas IO, Brau N, Waters M, et al. Tocilizumab in Hospitalized Patients with Severe Covid-19 Pneumonia. N Engl J Med 2021.

31. Hermine O, Mariette X, Tharaux PL, et al. Effect of Tocilizumab vs Usual Care in Adults Hospitalized With COVID-19 and Moderate or Severe Pneumonia: A Randomized Clinical Trial. JAMA Intern Med 2021;181:32–40.

32. Stone JH, Frigault MJ, Serling-Boyd NJ, et al. Efficacy of Tocilizumab in Patients Hospitalized with Covid-19. N Engl J Med 2020;383:2333–44.

33. Salama C, Han J, Yau L, et al. Tocilizumab in Patients Hospitalized with Covid-19 Pneumonia. N Engl J Med 2021;384:20–30.

34. Thompson RN, Hill EM, Gog JR. SARS-CoV-2 incidence and vaccine escape. Lancet Infect Dis 2021.

